# MRI-derived Articular Cartilage Strains Predict Patient-Reported Outcomes Six Months Post Anterior Cruciate Ligament Reconstruction

**DOI:** 10.1101/2024.04.27.24306484

**Authors:** Emily Y. Miller, Woowon Lee, Timothy Lowe, Hongtian Zhu, Pablo F. Argote, Danielle Dresdner, James Kelly, Rachel M. Frank, Eric McCarty, Jonathan Bravman, Daniel Stokes, Nancy C. Emery, Corey P. Neu

## Abstract

**Key terms:** Multicontrast and Multiparametric, Magnetic Resonance Imaging, Osteoarthritis, Functional Biomechanical Imaging, Knee Joint Degeneration

**What is known about the subject:** dualMRI has been used to quantify strains in a healthy human population *in vivo* and in cartilage explant models. Previously, OA severity, as determined by histology, has been positively correlated to increased shear and transverse strains in cartilage explants.

**What this study adds to existing knowledge:** This is the first *in vivo* use of dualMRI in a participant demographic post-ACL reconstruction and at risk for developing osteoarthritis. This study shows that dualMRI-derived strains are more significantly correlated with patient-reported outcomes than any MRI relaxometry metric.

**Background:** Anterior cruciate ligament (ACL) injuries lead to an increased risk of osteoarthritis, characterized by altered cartilage tissue structure and function. Displacements under applied loading by magnetic resonance imaging (dualMRI) is a novel MRI technique that can be used to quantify mechanical strain in cartilage while undergoing a physiological load.

**Purpose:** To determine if strains derived by dualMRI and relaxometry measures correlate with patient-reported outcomes at six months post unilateral ACL reconstruction.

**Study Design:** Cohort study

**Methods:** Quantitative MRI (T2, T2*, T1ρ) measurements and transverse, axial, and shear strains were quantified in the medial articular tibiofemoral cartilage of 35 participants at six-months post unilateral ACL reconstruction. The relationships between patient-reported outcomes (WOMAC, KOOS, MARS) and all qMRI relaxation times were quantified using general linear mixed-effects models. A combined best-fit multicontrast MRI model was then developed using backwards regression to determine the patient features and MRI metrics that are most predictive of patient-reported outcome scores.

**Results:** Higher femoral strains were significantly correlated with worse patient-reported functional outcomes. Femoral shear and transverse strains were positively correlated with six-month KOOS and WOMAC scores, after controlling for covariates. No relaxometry measures were correlated with patient-reported outcome scores. We identified the best-fit model for predicting WOMAC score using multiple MRI measures and patient-specific information, including sex, age, graft type, femoral transverse strain, femoral axial strain, and femoral shear strain. The best-fit model significantly predicted WOMAC score (p<0.001) better than any one individual MRI metric alone. When we regressed the model-predicted WOMAC scores against the patient-reported WOMAC scores, we found that our model achieved a goodness of fit exceeding 0.52.

**Conclusions:** This work presents the first use of dualMRI *in vivo* in a cohort of participants at risk for developing osteoarthritis. Our results indicate that both shear and transverse strains are highly correlated with patient-reported outcome severity could serve as novel imaging biomarkers to predict the development of osteoarthritis.

## Introduction

Anterior cruciate ligament (ACL) injuries are among the most common knee injuries, with more than 250,000 treated in the U.S. annually ^20^. Although ACL-reconstructive (ACLR) surgery consistently improves joint mechanics, patients display a substantial increased risk of developing knee osteoarthritis (OA) ^27,46,49^. Twenty-seven percent of individuals develop radiographic OA within five years of surgical ACLR, and upwards of 50%-90% develop OA within 10-20 years ^38^. Previous work has shown cartilage compositional changes occur as early as six months post-ACLR, yet symptomatic and radiographic cartilage changes do not manifest until 5-10 years post-ACLR ^34,46^. Furthermore, diagnosis often occurs after irreversible changes to the joint have occurred, resulting in an osteotomy or total knee arthroplasty as the only palliative options ^44^. Although the onset of OA after ACL injury happens early and its prevalence is high, the understanding of mechanisms initiating the degeneration of articular cartilage is limited. Therefore, the development of methods to understand and assess the degenerative changes that precede symptomatic OA is necessary to optimize interventions that may prevent future disease onset.

Magnetic resonance imaging (MRI) is a promising imaging modality for the clinical assessment of preradiographic OA cartilage changes such as cartilage thinning, and cartilage compositional changes. Multiple semi-quantitative scoring systems have been introduced that evaluate cartilage damage, subchondral bone lesions, subchondral cysts, and other morphological features ^23–26,40,42^. Notwithstanding the significance of these scoring systems, the early degradation of the cartilage tissue matrix is not visible with these methods. Quantitative MRI (qMRI) relaxometry maps (i.e., T2, T2*, and T1ρ), have been developed that can bridge this gap by assessing collagen content, water content, and macromolecule organization^17,33,37,43,47,48^. Recently, as a mechanical complement to qMRI, we pioneered displacement under applied loading MRI (dualMRI) to measure cartilage mechanical properties and quantify joint health ^6,31,32,55^. dualMRI quantifies pixel-level full-field displacement and strain maps (i.e., “elastography” maps^4,29^) of cartilage while under a cyclical physiological load within the imaging scanner, mimicking a walking cadence. dualMRI represents a direct, image-based mechanics approach for measuring cartilage deformation under load noninvasively. We discovered that dualMRI is robust to quantify strain in silicone phantoms, bovine explant studies, and *in vivo* human participants ^31,32^. Moreover, compared to qMRI, dualMRI quantified shear strains in human cartilage explants more strongly correlate with histologically-assessed OA severity ^19^.

In this work, we have applied dualMRI and qMRI techniques in a clinical cohort of patients six months post-ACLR to determine which MRI metrics may best predict patient-reported outcomes. While previous work has quantified macro-scale whole joint mechanical changes post ACLR^3,18,21,50^, to our knowledge, this is the first study that has quantified cartilage tissue-level (i.e., intratissue) mechanical changes in this clinical population. This study had two objectives: first, we evaluated the extent that dualMRI and qMRI metrics are associated with knee function and pain at six-months post-ACLR, as assessed by standardized patient-reported outcome scores ^16,41^. Based upon our prior work in cartilage explants, we hypothesize that increased shear and transverse strains will be associated with worse functional outcomes as determined by patient-reported outcome scores. Second, we tested whether a multiparametric approach improves prediction of patient-reported outcome severity compared to any single MRI method alone ^19^. For this second objective, we hypothesize that a combined “best-fit” predictive model containing dualMRI, qMRI, and patient information will better predict patient-reported outcomes than any single MRI measure alone.

## Methods

We collected multicontrast MRI data on participants who underwent unilateral ACLR surgery six months prior to the scan date. We then examined the relationship between individual MRI metrics (transverse strain, shear strain, axial strain, T2, T2*, and T1ρ) and patient-reported outcome scores.

### Participants

A cross-sectional study design, which included participants who were recruited into a larger prospective longitudinal study, was used. All participants provided informed consent and were recruited and evaluated between 2022 and 2023 in accordance with Institutional Review Board approval at the University of Colorado. Participants were initially identified and recruited in the orthopedic clinic following unilateral ACL reconstructive surgery (Figure 1). All participants had surgery within three months of the initial injury, no prior history of concomitant, symptomatic knee pathologies on their ACL reconstructed knee and no injury or surgery on their contralateral knee. No participants were younger than age 18 or older than age 40. Participants were excluded if they had received a prior ACL reconstructive surgery on either limb, revision surgery, or any cartilage chondral defect surgery (Figure 1). All participants received either a bone-patella tendon-bone autograft or quadriceps tendon autograft at the recommendation of one of three collaborating orthopedic surgeons at University of Colorado Anschutz. Rehabilitation was standardized across the cohort based upon the MOON ACL rehabilitation protocol ^54^. At the time of the MRI scan, all participants completed the Knee Injury and Osteoarthritis Outcome Score (KOOS), Western Ontario and McMaster Universities Osteoarthritis Index (WOMAC), and Marx Activity Rating (MARS) questionnaires.

**Figure 1:**
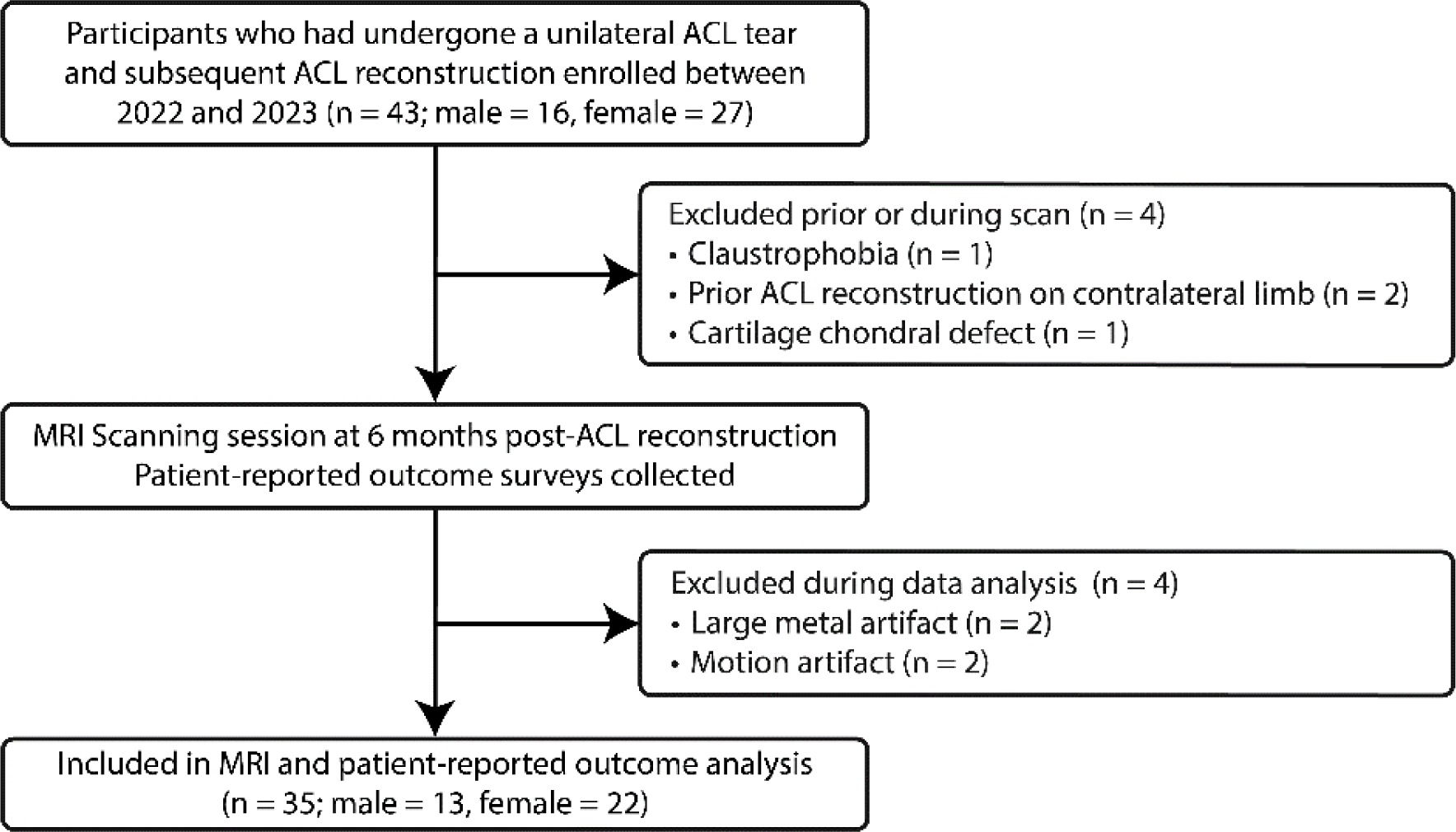
Flow diagram of participant selection and inclusion criteria. The experimental design included recruiting in the orthopedic clinic, 43 participants who had received a unilateral ACL reconstruction. Four participants were excluded prior to the MRI scan due to claustrophobia, prior ACLR on the contralateral limb, or cartilage chondral defect. After scanning, four more participants were excluded from analysis due to the presence of large metal artifact in the cartilage region of interest, or motion artifact that obscured analysis. ACL, anterior cruciate ligament. MRI, magnetic resonance imaging.

### Loading Protocol of Human Knee

An MRI-compatible loading apparatus capable of providing a varus load to the tibiofemoral cartilage of the knee, leading to compression of the medial femoral condyle, was used (Figure 2) ^55^. Moment conservation was used to calculate the load applied on the foot to be equivalent to 0.5 times body weight of load applied at the medial condyle. Patients were subjected to a 0.5Hz cyclic loading regime (i.e., pneumatic actuation, 1s load,1s unload) to mimic a walking cadence^32,55^.

**Figure 2:**
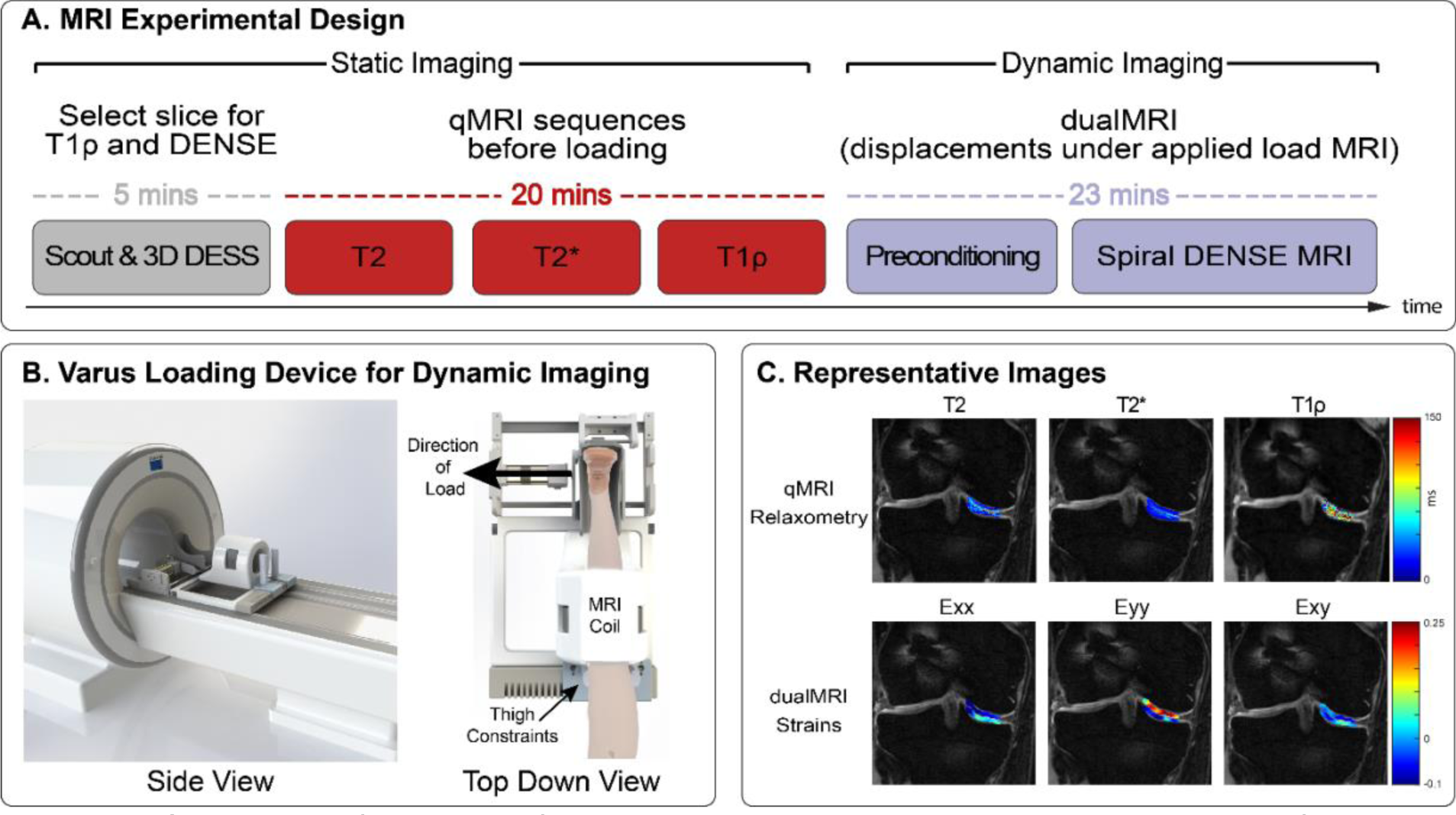
Schematic of MRI scan for each participant. A. The experimental design for acquiring qMRI relaxometry values (T2, T2*, T1ρ) during rest, and spiral displacement encoding with stimulated echoes (DENSE) MRI images during varus loading. First, a scout and 3D (double echo steady state (DESS) sequences were run to visualize knee anatomy. A slice was then selected where tibiofemoral contact in the medial condyle was maximized. Following DESS, relaxometry measurements (T2, T2*, T1ρ) were then acquired. B. We then implemented an MRI-compatible varus loading device, consisting of a thigh constraint, pneumatic cylinder, and boot section. Varus load was applied in a cyclic fashion consisting of 1 s of load followed by 1 s of unload. Preconditioning was applied for 8 minutes prior to DENSE acquisition to minimize the effect of cartilage viscoelasticity and motion artifacts. C. Following DENSE acquisition, phase images in *x* and *y* were converted to displacement fields in the selected cartilage regions of interest, smoothed, and then used to calculate Green-Lagrange strains. All participants were positioned in a supine position inside the MRI bore during all sequences. DESS, Double echo steady state. DENSE, Displacement encoding with stimulated echoes. qMRI, quantitative magnetic resonance imaging.

### MRI Scanning Protocol

The knee imaging protocol consisted of a fast gradient echo MR image sequence (scout) for localization and subsequently 3D double echo steady state (DESS) acquisition, quantitative T2, T2*, and T1ρ measurements, and a displacement encoded with stimulated echoes (DENSE) sequence acquired during cyclic varus loading on the knee joint (Figure 2) ^28,31,32^. All imaging was performed using a clinical MRI system (3 T; Siemens Magneton Prisma^fit^) with a 15-channel quadrature knee coil (Tx/Rx Knee 15 Flare Coil, QED, LLC). To minimize the viscoelastic response and motion artifact, the cartilage was preconditioned to reach a quasi-steady-state load-displacement response ^5–9,31,32^ by pre-applying eight minutes of cyclic loading prior to image acquisition using spiral DENSE MRI acquisition ^26,27^. Ten spiral interleaves were used to collect k-space data with a temporal resolution was 40 ms. The displacement encoding gradient was 0.64 cycles/mm. Other imaging parameters are listed in Table 1.

**Table 1:** MRI scanning parameters^a^.

|  | ***T1ρ*** | ***T2*** | ***T2\**** | ***DENSE*** |
| --- | --- | --- | --- | --- |
| ***TE (ms)*** | 3 ms | 13 ms | 4.87 ms | 2.5 ms |
| ***Echo train length*** | 4 | 6 | 8 | N/A |
| ***TR (ms)*** | 6 | 1290 | 1270 | 20 |
| ***FOV (mm<sup>2</sup>)*** | 100x100 | 100x100 | 100x100 | 160x160 |
| ***Spatial resolution (μm<sup>2</sup>)*** | 700x700 | 390x390 | 390x390 | 340x340 |
| ***Slice thickness (mm)*** | 3.5 | 1.7 | 2 | 1.7 |
| ***Number of slices*** | 1 | 13 | 11 | 1 |
| ***Flip angle (°)*** | 10 | 180 | 60 | 20 |
| ***Averages*** | 2 | 2 | 1 | 8 |
| ***Acquisition time (min)*** | 11 | 10 | 9 | 14 |
| ***Fit Type*** | Mono-exponential relaxation model<sup>2,51</sup> | Mono-exponential using non-negative least squares<sup>35,52</sup> | Mono-exponential using non-negative least squares<sup>35,39,52</sup> | N/A |

<sup>a</sup> DENSE, Displacement encoded with stimulated echoes.

### MRI Data Analysis

The articular cartilage was semi-automatically segmented into two regions of interest (ROI) based on the anatomical regions of the knee (i.e., medial femoral cartilage, medial tibial cartilage). Cartilage segmentation was repeated on all 27 time frames. Locally weighted scatterplot smoothing (LOWESS) was applied to the raw displacement within each ROI using a span of 10 for 30 cycles. In-plane Green-Lagrange strains (axial, transverse, and shear) were calculated in the ROIs from the smoothed displacements using the deformation gradient tensor ^7–9,31,32^. All left limb scans were converted to a right limb coordinate system to facilitate strain comparisons. To provide a single measure that incorporates all three finite strains, Von Mises strains (VMS) were calculated as previously described ^19^. On the T2, T2*, and T1ρ maps, ROIs were selected that were analogous to the medial cartilage tibial and femoral DENSE ROIs. The mean value for each MRI metric (transverse strain, axial strain, shear strain, VMS, T2, T2*, and T1ρ) within each ROI (medial tibial and medial femoral) was then calculated for statistical analysis. Sample maps of all MRI metrics are shown in Figure 2.

### Statistical analysis

All statistical analyses were performed with SAS Version 9.4 (SAS Institute), with a 2-sided significance level of ɑ=0.05. To evaluate the effects of patient factors on each MRI metric, we created a separate linear mixed effects model for each MRI response variable. Sex, surgeon, graft type, and location were fixed categorical effects, while the age, BMI, and days from surgery were covariates. Patient ID was a random effect. We evaluated the residuals from each model to confirm they met the assumptions of ANOVA^19^. Any data that did not meet the assumption of normality as determined by the Shapiro-Wilk test was logarithmically transformed using standard methods. We then evaluated the relationships between patient-reported outcomes and MRI metrics using general linear models. A separate model was developed for each of the two patient-reported outcomes (KOOS and WOMAC). In each model, sex, surgeon, and graft type were fixed categorical effects, and age, BMI, and days from surgery were covariates. We again evaluated the residuals from each model to test for normality and homoskedasticity of residuals. We identified the best-fit model for predicting patient-reporting outcomes using backward regression. We started with a full model that included all 12 MRI metrics as well as sex, age, surgeon, graft type, BMI, and days from surgery as predictor variables. A separate backward regression was developed for each patient-report outcome. The best-fit model was selected from the initial pool of potential variables using AIC minimization. Finally, we evaluated the extent to which the best-fit models predicted each patient-reported outcome using simple linear regression.

## Results

### Participant Characteristics

The average participant age was 25.9±5.8 and mean body mass index (BMI) was 24.2±4.1. The participant demographic and surgical features are shown in Table 2.

**Table 2:** Participant demographic, surgical characteristics, and patient-reported outcomes^a^.

| Variable | Value |
| --- | --- |
| Age at scan date (years) | 25.9 ± 5.8 |
| BMI (kg/m<sup>2</sup>) | 24.2 ± 4.1 |
| Time since ACLR at MRI Scan (months) | 6.8 ± 1.1 |
| Sex |  |
| Male | 13 (37.1%) |
| Female | 22 (62.8%) |
| Graft Type |  |
| Bone-patellar tendon-bone autograft | 28 (80%) |
| Quadriceps tendon autograft | 7 (20%) |
| KOOS Score | 74.3 ± 10.1 |
| Symptoms | 81.7 ± 12.1 |
| Pain | 85.0 ± 8.8 |
| Activities of Daily Living | 92.9 ± 9.5 |
| Sports and Recreation | 60.1 ± 18.5 |
| Quality of Life | 51.4 ± 51.4 |
| WOMAC Score | 7.8 ± 7.6 |
| Pain | 1.2 ± 1.5 |
| Stiffness | 1.6 ± 1.3 |
| Physical Function | 4.9 ± 5.6 |
| MARS Score | 7.9 ± 5.1 |

<sup>a</sup>Data are reported as mean ± SD, or n (%). All KOOS scores are on a 0-100 scale, with 0 representing extreme knee problems. All WOMAC scores are on a scale of 0 to 96, with 96 representing extreme knee problems. WOMAC subscore pain is scored from 0-20, stiffness from 0 to 8, and physical function from 0 to 68. BMI, body mass index. ACLR, anterior cruciate ligament reconstruction. KOOS, Knee Injury and Osteoarthritis Outcome Score. WOMAC, Western Ontario and McMaster Universities Osteoarthritis Index, MARS, Marx Activity Rating (MARS)

### Relationships between MRI metrics and Patient Factors

There were statistically significant (p<0.05) associations between shear strain and graft type, with shear strains in participants who had received a bone-patellar-tendon bone graft significantly higher than those who had received a quadriceps tendon graft. (p = 0.03, Table 3). Average T2 values were significantly lower in the quadriceps tendon group (p=0.01, Table 3), as well as significantly lower on average in female participants (p=0.006, Table 3). Sex differences were also observed in average T2* values, with female participants displaying significantly lower average T2* values (p=0.003, Table 3). Both T1ρ and T2* values were significantly lower in the tibial ROI than in the femoral ROI (p < 0.001 and p < 0.001, respectively, Table 3).

**Table 3:** Linear mixed-model estimates for associations between MRI metrics and patient factors^a^.

| Term | Level | Transverse Strain | Axial Strain | Shear Strain | Von Mises Strain | T2 (ms) | T1ρ (ms) | T2* (ms) |
| --- | --- | --- | --- | --- | --- | --- | --- | --- |
| Sex | Female | 0.016<br>(0.010, 0.023) | -0.017<br>(-0.041, 0.007) | 0.019<br>(0.011, 0.027) | 0.111<br>(0.091, 0.132) | 35.9<br>(32.1, 39.6) | 25.0<br>(23.5, 26.5) | 17.8<br>(15.8, 19.7) |
|  | Male | 0.005<br>(-0.005, 0.014) | -0.008<br>(-0.041, 0.026) | 0.008<br>(-0.002, 0.019) | 0.097<br>(0.067, 0.126) | 44.1<br>(39.1, 49.0) | 25.5<br>(23.3, 27.6) | 24.3<br>(21.5, 27.1) |
|  | Significance | p=0.07 | p=0.67 | p=0.13 | p=0.29 | **p=0.006** | p=0.75 | **p=0.003** |
| Graft Type | Patellar | 0.014<br>(0.008, 0.020) | -0.009<br>(-0.028, 0.0098) | 0.018<br>(0.012, 0.025) | 0.113<br>(0.097, 0.130) | 37.1<br>(33.9, 40.3) | 25.2<br>(24.1, 26.5) | 19.8<br>(18.2, 21.3) |
|  | Quadriceps | 0.002<br>(-0.008, 0.014) | -0.031<br>(-0.073, 0.009) | 0.003<br>(-0.010, 0.016) | 0.078<br>(0.043, 0.113) | 46.1<br>(40.3, 51.9) | 24.7<br>(22.1, 27.4) | 21.9<br>(18.5, 25.2) |
|  | Significance | p=0.06 | p=0.32 | **p=0.03** | p=0.07 | **p=0.01** | p=0.72 | p=0.27 |
| Location | Femoral | 0.015<br>(0.006, 0.024) | -0.001<br>(-0.024, 0.021) | 0.020<br>(0.009, 0.031) | 0.109<br>(0.089, 0.130) | 39.3<br>(36.8, 41.8) | 27.7<br>(26.3, 29.1) | 23.0<br>(21.5, 24.4) |
|  | Tibial | 0.009<br>(0.005, 0.015) | -0.026<br>(-0.049, 0.003) | 0.010<br>(0.005, 0.016) | 0.103<br>(0.085, 0.120) | 38.6<br>(33.3, 43.9) | 22.6<br>(21.3, 24.0) | 17.4<br>(15.9, 19.0) |
|  | Significance | p=0.31 | p=0.11 | p=0.15 | p=0.70 | p=0.14 | **p<0.001** | **p<0.001** |

### Correlation of MRI metrics and 6-month patient-reported outcome scores

Higher femoral strains were correlated with worse functional outcomes. Femoral shear and transverse strains were positively correlated with 6-month WOMAC scores (Figure 3). After controlling for covariate and fixed categorical effect data, the strength of the correlations with the WOMAC score increased for both transverse and shear strain (transverse: p < 0.001, shear: p = 0.006, Figure 3). Similarly, the unadjusted shear strains were significantly correlated with KOOS scores (Figure S1, supplement). When adjusted for covariate data, the strength of the shear strain correlation increased (p = 0.01), and both transverse and Von Mises strain became significant (transverse: p = 0.02, Von Mises: p = 0.04). In general, qMRI measures related poorly to both KOOS and WOMAC scores.

**Figure 3:**
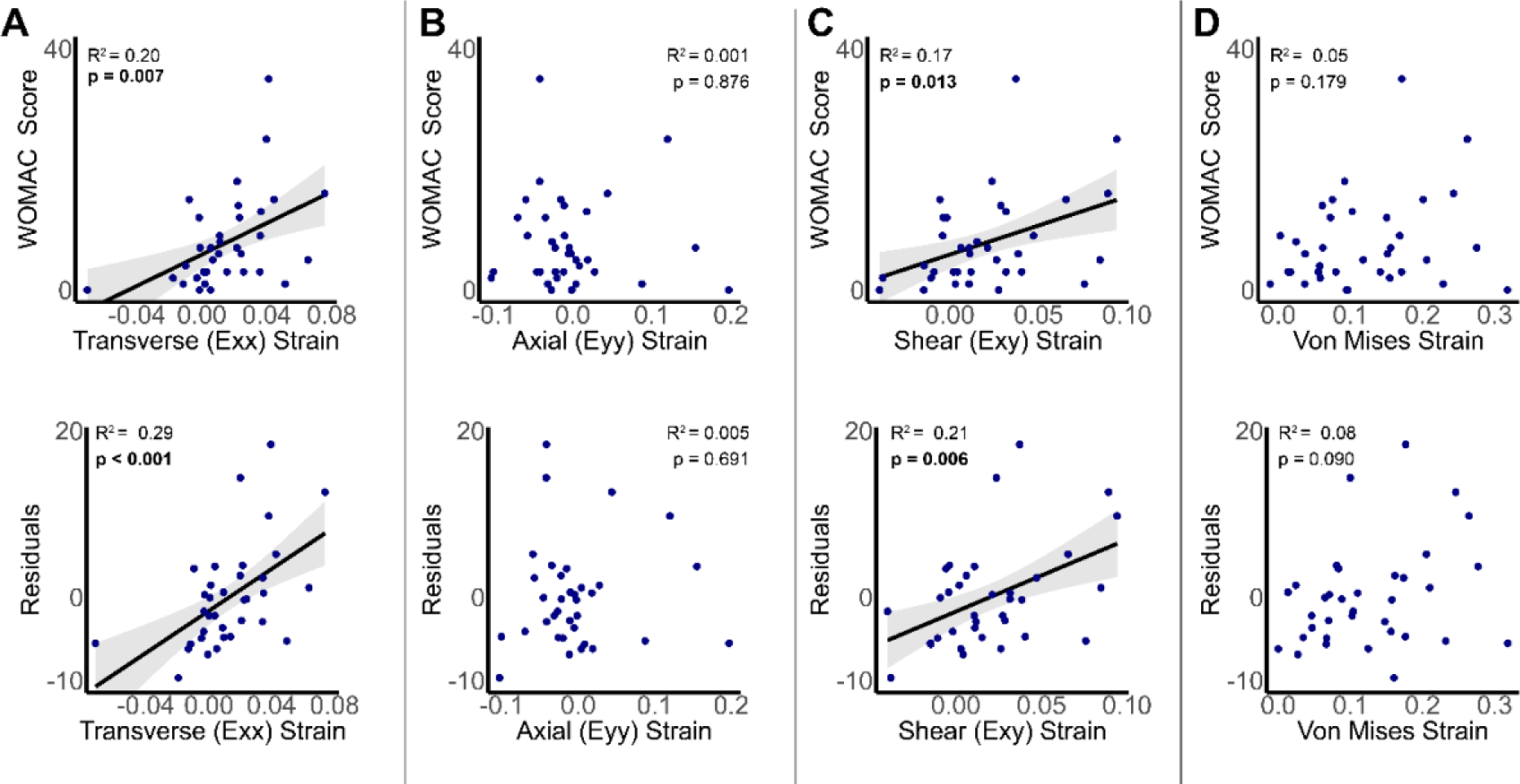
Femoral strain measures are significantly correlated with patient reported outcome severity. Unadjusted transverse (A, top row) and shear (C, top row) strains significantly correlate with WOMAC score (left), whereas axial strains (B) and Von Mises (D) strains were variable. After controlling for variability introduced by covariate data such as graft type, age, BMI, sex, and time since surgery, the correlation for both transverse and shear strains improved (A, C, bottom row). Correlations with the KOOS scores are shown in Supplement S1. All tibial data are shown in supplement S4 and S%. Confidence intervals of 95% are shown in gray.

Additionally, unlike what was seen in strain values, controlling for covariates did not improve correlations between any qMRI relaxometry measure and any patient-reported outcome (Figure 4, Figure S2). No MRI measures of either dualMRI or qMRI in the tibial ROI were significantly correlated with either KOOS or WOMAC scores (Supplement, S4, S5). Furthermore, there were no correlations found between any MRI metric and MARS.

**Figure 4:**
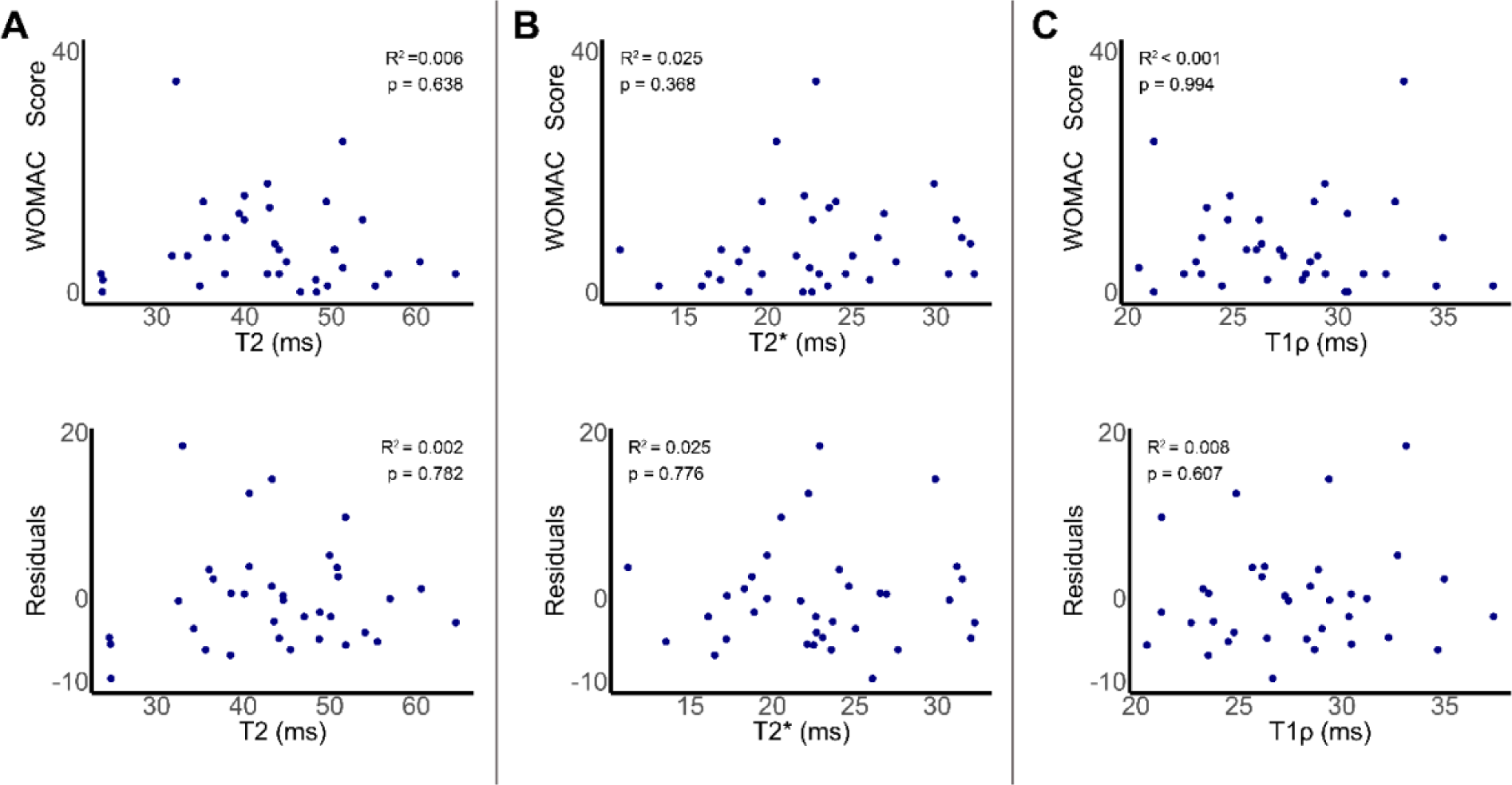
qMRI relaxometry (T2, T2*, T1ρ) measures did not significantly correlate with WOMAC scores. After controlling for potential covariates, trends generally worsened, although T1ρ (C) improved. All tibial correlations and correlations with KOOS score are shown in supplemental figures S2, S4, and S5.

### Multicontrast MRI Model

We identified the best-fit model for predicting WOMAC scores from MRI data and patient demographic information using backward regression. The best-fit model predicted WOMAC scores better than any one individual MRI metric alone. When we regressed the model-predicted WOMAC scores against the patient-reported WOMAC scores, we found that our model achieved high significance (p< 0.001, R^2^=0.52). The best-fit model for WOMAC scores contained sex, age, graft type, femoral transverse strain, femoral axial strain, and femoral shear strain (Figure 5). No tibial MRI metrics were included in the model. The best-fit model for KOOS contained the same variables as the WOMAC model with the addition of femoral T1ρ (Figure S3).

**Figure 5:**
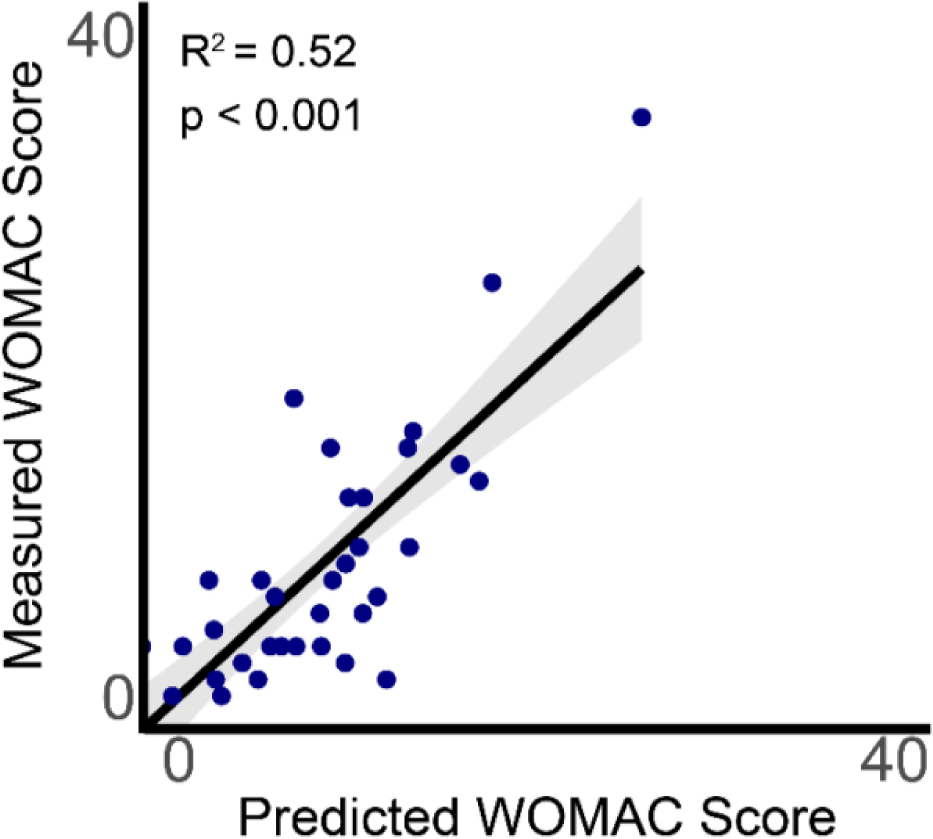
Multicontrast MRI measures best predict WOMAC score. The predicted WOMAC scores from a best-fit model that contains sex, age, graft type, femoral shear strain, femoral axial strain, and femoral transverse strain are shown on the x-axis, while the patient-reported WOMAC score is shown on the y-axis. Confidence intervals of 95% are shown in gray. [Mention/reference here also supplemental figure for similar response for KOOS]

## Discussion

A non-invasive method to accurately assess cartilage degeneration at its earliest stages is critical for the monitoring of the pre-osteoarthritic disease state. Quantitative evaluations of cartilage mechanical and biochemical properties could provide an early warning of pre-osteoarthritic cartilage changes that could be reflective of long-term joint health ^47^. This study aimed to utilize dualMRI and qMRI to evaluate early cartilage changes in a cohort of participants six months post-ACL reconstruction. Here we demonstrated three main findings: 1: Shear and transverse femoral strains were significantly correlated with patient-reported outcome scores, with higher strains indicating worse pain, stiffness, and functional limitations, 2: qMRI relaxometry techniques were not correlated with patient-reported outcome scores in this population, and 3: a combined “best-fit” multicontrast MRI model better predicted patient-reported outcome than any single MRI technique alone.

The findings of this study align with a previous study from our group that used dualMRI and qMRI in cartilage explants with histologically defined OA^19^. In our previous study, it was demonstrated that dualMRI strains (Von mises strain, transverse, and shear strain) were positively correlated with OA severity, as defined by histological assessment and the OARSI scoring system^1^. qMRI measures (including T2 and T1ρ scans) were also taken, and only T1ρ was found to be significantly correlated with the OARSI score^19^. Although in this study we did not find any significant correlations between T1ρ values and patient-reported outcomes, it is possible that six months post-ACL reconstruction is too early to detect significant cartilage matrix changes with T1ρ. This is supported by previous work that took T1ρ scans at baseline (after ACL injury but prior to reconstruction), six months, and one-year time points post-ACL reconstruction^43^. In this previous work, it was found that while T1ρ values were highly correlated with KOOS scores at both baseline and one-year time points, at the six-month timepoint, only the KOOS subscore for pain was significantly correlated with T1ρ values. Our study is also supported by another study that indicates that there is a weak association between loss of tibial cartilage and worsening of OA symptoms^53^. The translation of dualMRI from cartilage explants in previous work to the current *in vivo* human study of cartilage degeneration represents considerable technological development in bringing the dualMRI technique out of the laboratory and into the clinic. The fact that increased transverse and shear strains closely correlate with worse outcome measures of joint health in both explants and *in vivo* models, serves as further evidence of dualMRI’s clinical relevance as a technique to quantify cartilage degeneration.

Our study supports previous work that has found that mechanical metrics may predict osteoarthritis better than relaxometry measures^15^. In explant work that quantified cartilage creep and stiffness as well as T1ρ relaxation time, it was found that bivariate models using mechanical metrics showed an improved ability in predicting OA compared to models using only T1ρ relaxation time^14^. Other *in vivo* work has quantified axial strain in tibiofemoral cartilage by measuring cartilage thickness changes before and after load using morphological MRI, and then quantifying localized strain as the normalized difference in cartilage thickness^30^. It is possible that we may not have seen a relationship between axial strain and patient-reported outcomes, due to the low magnitude of load applied to the cartilage. Our loading device applied a load of one-half body weight to the medial cartilage, whereas other experiments have calculated axial strain based on cartilage thickness changes after significant physiological loads such as long-distance running or walking, or due to activities of daily living outside of the MRI scanner ^11–13,22,30^. In these specific situations, the cartilage would likely be loaded to at least twice body weight, and possibly as high as eight times body weight ^36^. Our work expands on the capabilities of MRI to quantify strain, as dualMRI quantifies intratissue strains with full complexity (i.e., shear, transverse, and axial strain). Here, we show that for predictive power of early degenerative cartilage changes, the full strain field is required. Additionally, this study positions dualMRI as an analysis tool of deformation measures that occur in more complex loading situations in other tissues (e.g., intervertebral disk, hip, and wrist). Multicontrast MRI, the combination of multiple MRI values in a model used to predict one patient-reported outcome, provided the strongest correlation with outcome severity. This is also supported by our group’s prior cartilage explant work^19^. It should be noted that our current model underpredicts worse functional outcomes. This work would benefit from the addition of other participants who score worse on patient-reported outcomes.

Our study had several limitations. The relatively small sample size limits the power of this work to fully comment on etiological factors such as surgical technique, graft tendon origin, or other demographic factors. However, the use of linear mixed effect models did allow us to account for all fixed effects and covariate data. Additionally, only average values for all MRI data were taken. It should be noted that splitting the cartilage into further sub-regions of interest (i.e., superficial vs deep zones), could be a subject of further study. Furthermore, while all participants were subject to the MOON ACL rehabilitation protocol, data was not collected on rehabilitation frequency or adherence. These factors all contribute to the variability between individuals and the mechanical loads that are experienced in their cartilage. Despite this, correlations of all MRI measures against the MARS score were insignificant. It is possible that this reflects an intentional limitation of activities until the 9-12 months post-surgical timepoint based upon rehab and return to play guidelines; however, it could also indicate that solely the act of taking part in activities that place high mechanical load demands on the knee joint may not be associated with altered cartilage composition as determined by qMRI. However, additional work has suggested that whole joint mechanical factors, including loss of knee extension and excessive tibial rotation, are associated with articular cartilage changes post-ACL reconstruction ^48^. This supports that abnormal loading, but not the intensity or frequency of load, could be a contributing cause of altered cartilage matrix changes. Further work comparing dualMRI-derived strains to results from kinematic gait analyses would provide further insight into cartilage degeneration as it relates to whole joint loading patterns and physical function ^32^. New technologies facilitating the use of markerless motion capture outside of a traditional gait biomechanics lab, may facilitate the ease of incorporating motion analyses into MRI studies ^45^.

Genetic and epidemiological studies have indicated that pre-OA disease states may potentially be modified ^10^. If OA can be diagnosed before irreversible changes occur, extrinsic factors such as obesity and joint loading patterns that are known to increase OA risk, could act as potential disease modification targets. To determine participants who would most benefit from interventions, imaging biomarkers that can identify pre-osteoarthritic cartilage changes are required. This work demonstrates dualMRI’s potential, particularly in combination with other MRI metrics, as a possible imaging biomarker for the identification of pre-osteoarthritis. While this work was only conducted in participants six months post-ACL reconstruction, greater longitudinal follow-up studies, potentially at 2-, 5-, and 10-year intervals would further refine this method for the longitudinal evaluation of cartilage degeneration.

### Conclusion

Our work presents the first use of dualMRI in an *in vivo* cohort of participants at risk for developing osteoarthritis. Our results indicate that both shear and transverse strains are correlated with patient-reported outcome severity and could be predictive of the development of osteoarthritis, suggesting that shear and transverse strains may serve as ideal biomarkers for pre-OA. These studies also demonstrate the sensitivity of dualMRI to cartilage changes over a short period of time (6 months), during which morphological MRI would not show cartilage changes. Our work presents the first study of dualMRI in an initial ACL reconstructed cohort; and supports the robustness of dualMRI to be used in future longitudinal studies to assess cartilage degeneration.

## Data Availability

All data produced in the present study are available upon reasonable request to the authors.

## Acknowledgments

This work was supported by NIH grant 2 R01 AR063712 and AR063712S1 (CPN). The authors are thankful for MRI technical support from Teryn S. Wilkes at the Intermountain Neuroimaging Consortium located at University of Colorado Boulder.

## Supplemental Figures

**Figure S1:**
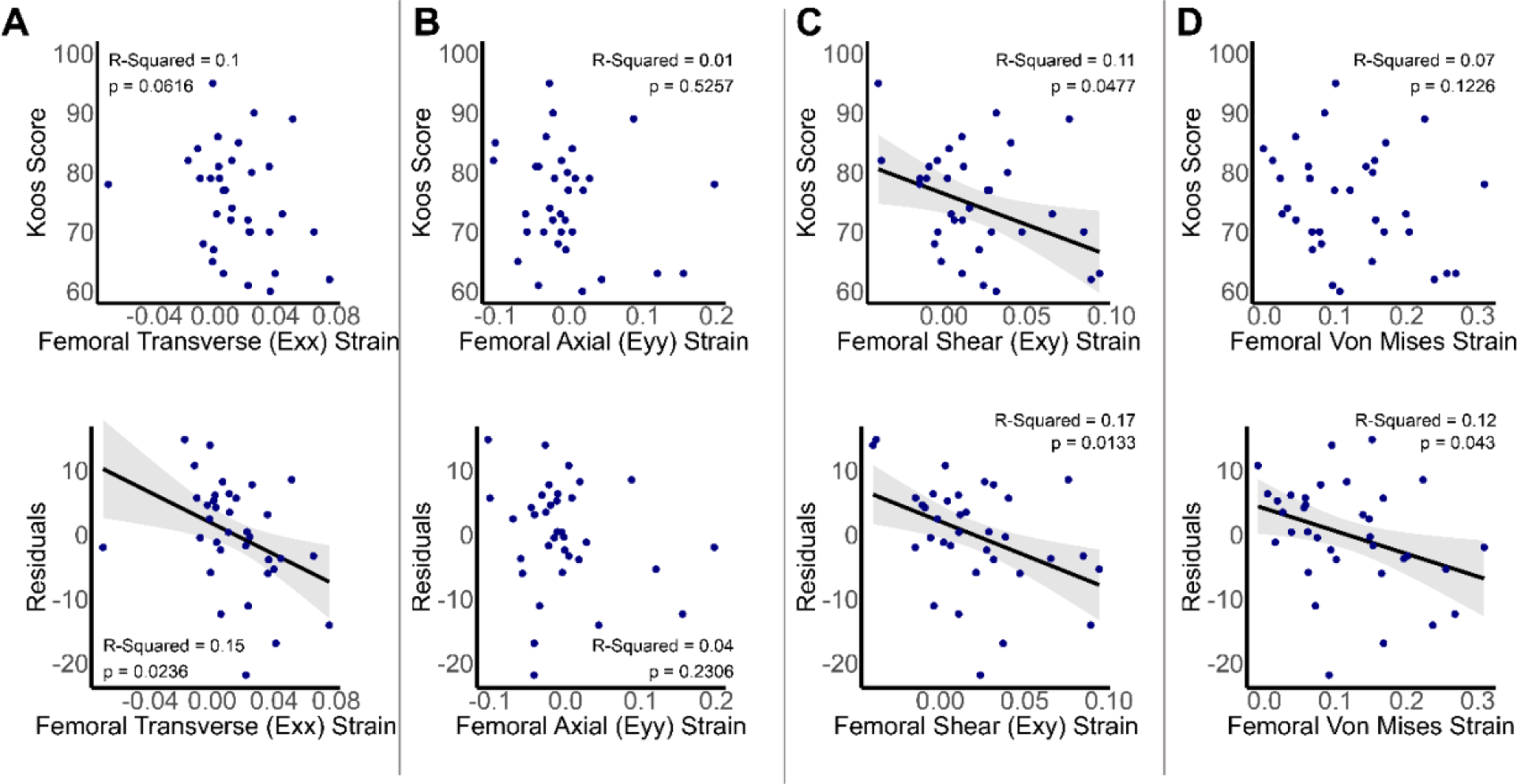
Femoral strain measures significantly correlate with patient-reported outcome severity. Unadjusted shear strains (top row) significantly correlate with KOOS score (left), whereas axial, transverse, and von mises strains are variable. After controlling for variability introduced by covariate data such as graft type, age, BMI, sex, and time since surgery, the correlation greatly improves, and the relationship between KOOS score and strain becomes significant for transverse and Von Mises strain (bottom row). Confidence intervals of 95% are shown in gray.

**Figure S2:**
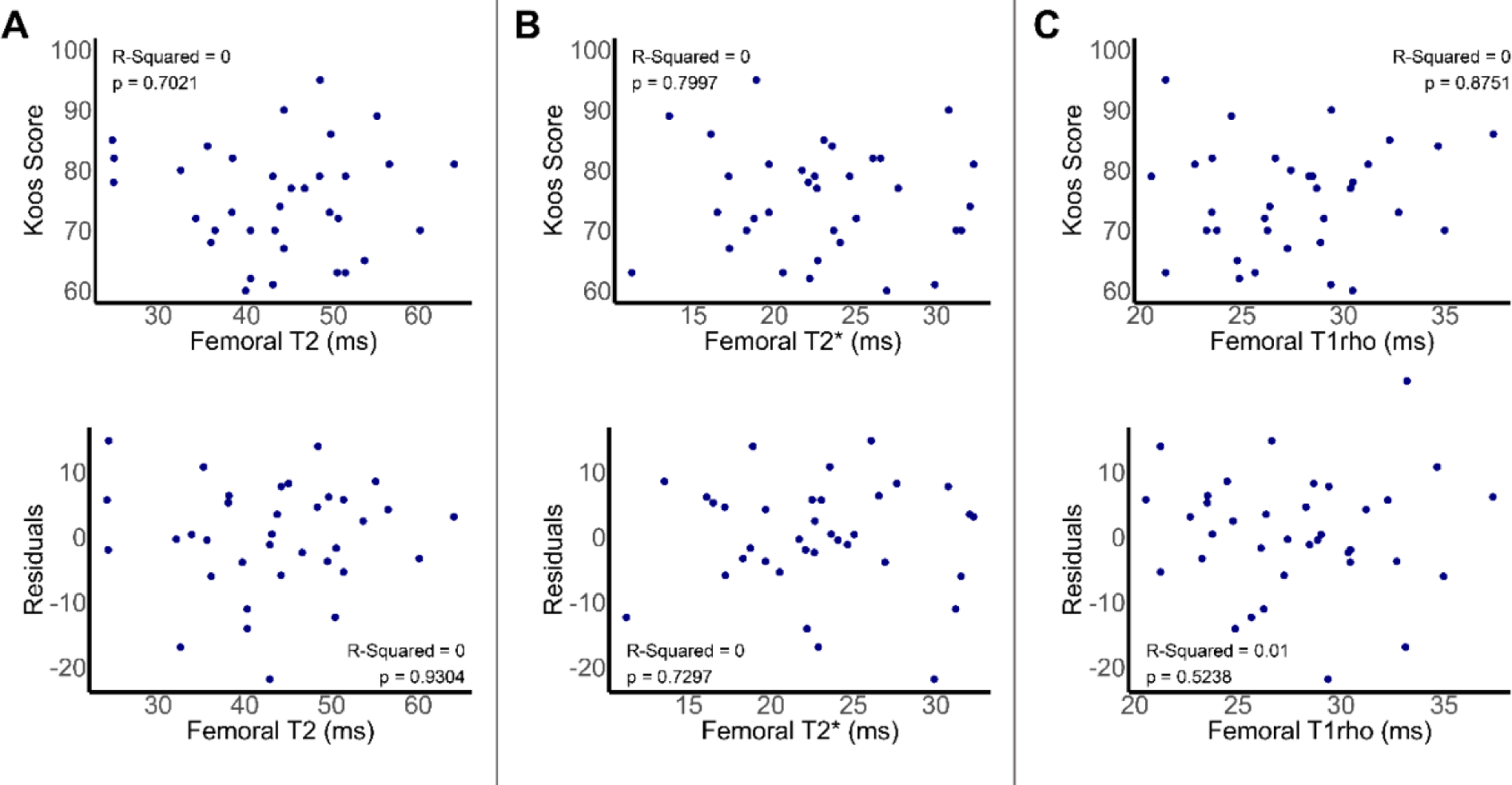
qMRI relaxometry measures do not significantly correlate with KOOS score. After controlling for potential covariates (bottom row), trends generally worsened, although T1ρ improved.

**Figure S3:**
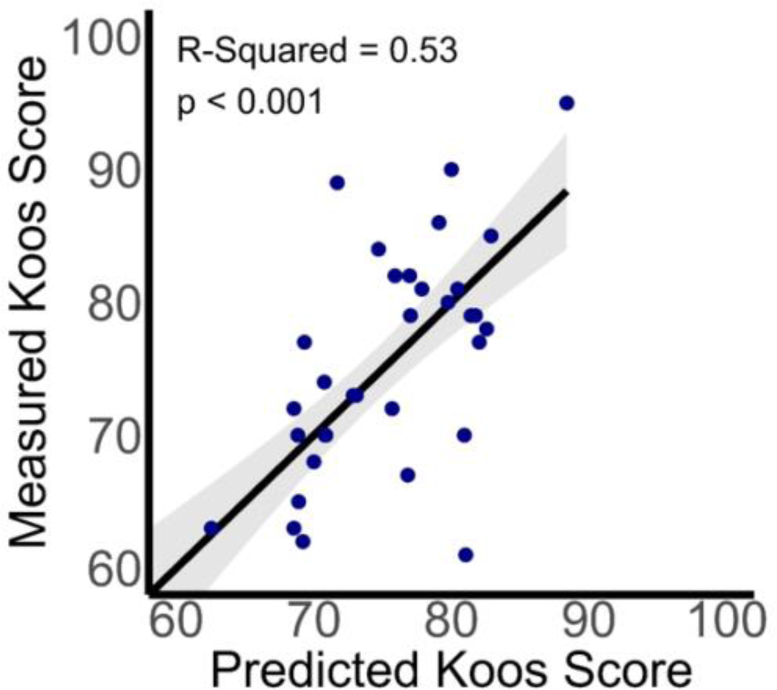
Multicontrast MRI measures best predict KOOS score. The predicted KOOS scores from a best-fit model that contains sex, age, graft type, femoral shear strain, femoral axial strain, and femoral transverse strain are shown on the horizontal axis, while the patient-reported KOOS score is shown on the vertical axis. Confidence intervals of 95% are shown in gray.

**Figure S4:**
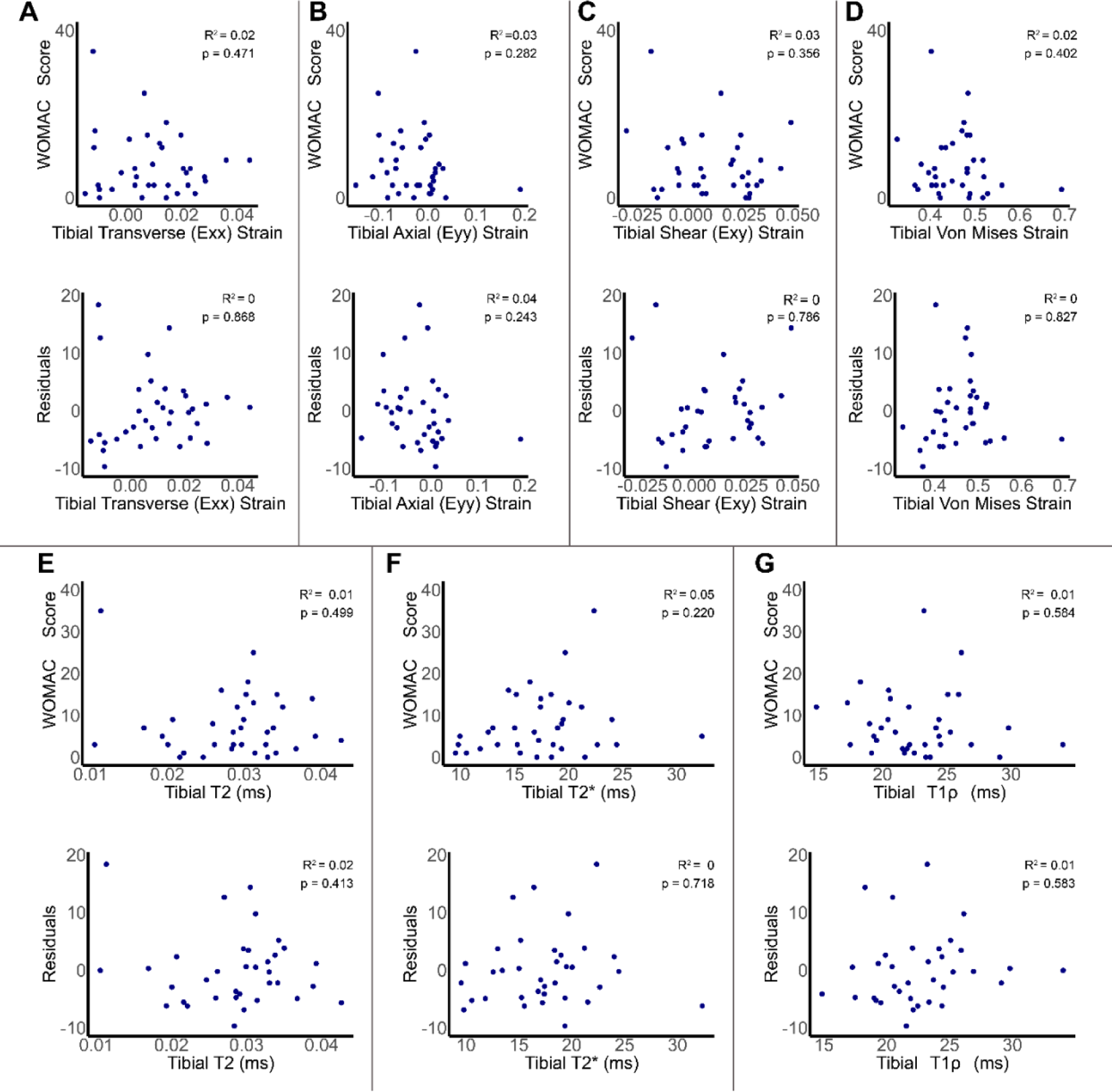
qMRI relaxometry measures and dualMRI derived strains in the tibial articular cartilage region of interest do not significantly correlate with WOMAC score.

**Figure S5:**
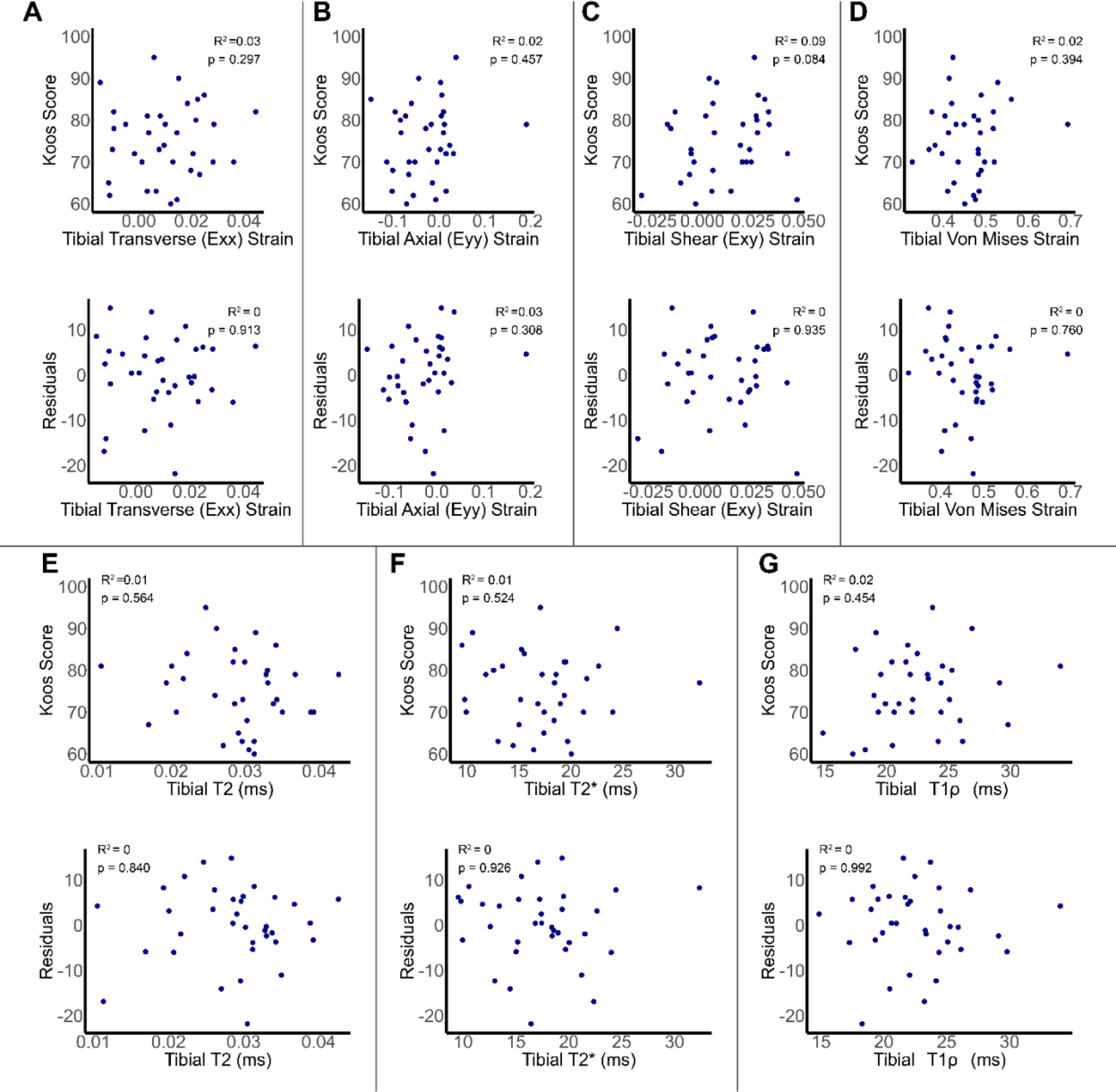
qMRI relaxometry measures and dualMRI derived strains in the tibial articular cartilage region of interest do not significantly correlate with KOOS score.

